# Preterm formula, fortified or unfortified human milk for very preterm infants, the PREMFOOD study, a parallel randomised feasibility trial

**DOI:** 10.1101/2023.10.31.23297886

**Authors:** Luke Mills, Karyn E Chappell, Robby Emsley, Afshin Alavi, Izabela Andrzejewska, Shalini Santhakumaran, Richard Nicholl, John Chang, Sabita Uthaya, Neena Modi

## Abstract

**Objective:** Uncertainty exists regarding optimal supplemental diet for very preterm infants if mother’s own milk (MM) is insufficient. We evaluated feasibility for a randomised controlled trial (RCT) powered to detect important differences in health outcomes.

**Methods:** In this open, parallel, feasibility trial, we randomised infants 25+0-31+6 weeks gestation by opt-out consent, to one of three diets: unfortified human milk (UHM) (unfortified MM and/or unfortified pasteurised human donor milk (DM) supplement; fortified human milk (FHM) (fortified MM and/or fortified DM supplement), and unfortified MM and/or preterm formula (PTF) supplement from birth to 35+0 weeks post menstrual age. Feasibility outcomes included opt-outs, adherence rates, and slow growth safety criteria. We also obtained anthropometry, and magnetic resonance imaging body composition data at term and term plus 6 weeks (opt-in consent).

**Results:** 35 infants were randomised to UHM, 34 to FHM, and 34 to PTF groups, of which 21, 19, and 24 infants completed imaging at term. Study entry opt-out rate was only 38%, while 6% of parents subsequently withdrew from feeding intervention. Two infants met predefined slow weight gain thresholds. There were no significant between-group differences in total adipose tissue volume at term (mean (sd): UHM: 0.870L (0.35L); FHM: 0.889L (0.31L); PTF: 0.809L (0.25L), p=0.66), nor in any other body composition measure or anthropometry at either timepoint.

**Conclusions:** Randomisation to UHM, FHM, and PTF feeding interventions by opt-out consent was acceptable to parents and clinical teams, associated with safe growth profiles and no significant differences in body composition. Our data provide justification to proceed to a larger RCT.

## Introduction

Uncertainty exists regarding the optimal supplement for very preterm infants when mother’s own milk (MM) is insufficient. Pasteurised human donor milk (DM) and preterm formula (PTF) are options. Unlike PTF, due to variable nutrient content of human milk, many practitioners also provide bovine-derived multicomponent fortification. Current meta-analyses of preterm feeding trials have found no difference in necrotising enterocolitis (NEC) with fortified versus unfortified human milk [1], and a higher risk of NEC with formula versus donor milk as sole feed [2]. However, interpretation is limited by inadequate power for NEC as a primary outcome, variable use of bovine fortifier, and type of formula. Although faster short-term growth was associated with formula or fortifier in these trials, there was no difference in long-term anthropometry, nor many other long-term health outcomes in the few trials that have evaluated for these [1,2]. Nevertheless, the increased risk of metabolic syndrome and cardiovascular disease [3–4] among very preterm survivors raises the possibility of early nutrition mediating these effects through rapid growth and/or altered body composition. Using whole body Magnetic Resonance Imaging (MRI), our group have previously shown that total and internal abdominal (visceral) adipose tissue (IAAT), a biomarker of long-term metabolic risk [4], is increased in young adults born preterm [5], with increases in IAAT seen as early as term equivalent age [6], compared with individuals born full term.

Clinical trials addressing these important uncertainties require sample sizes with adequate power to detect meaningful differences in important outcomes and establish cohorts for long-term follow up, with design safe and appropriate to maximise recruitment and completion of feed intervention.

The objectives of this feasibility trial were to assess (1) feeding intervention opt-out consent and adherence rates (2) trial DM volume requirement (3) anthropometry and MRI body composition at term and term plus 6 weeks, and (4) to test that clinical outcome data can be retrieved from a national database of real-world clinical data.

## Materials and Methods

We conducted PREMFOOD (PREterM FOrmula Or Donor milk for preterm babies), an open, multicentre, parallel, feasibility randomised controlled trial (RCT), pre-registered at ClinicalTrials.gov (NCT01686477) and approved by the UK National Research Ethics Service (REC no: 12/LO/1391). Recruitment occurred from August 2013 to November 2017, at three London National Health Service hospitals.

Preterm infants born between 25+0 and 31+6 weeks who were unlikely to be transferred to another hospital, were eligible for inclusion. Infants with congenital abnormalities that precluded early milk feeding were excluded. The study was discussed with parents at the time of birth, with opportunity offered to opt-out; parents were informed that they had the right to change a decision to participate at any time. Subsequent randomisation to feed intervention occurred postnatally within 48hrs unless parents opted-out. Opt-out is a valid, ethical method for informed consent in neonatal research [7] and was chosen for the comparative-effective component of the trial as this reduces the burden of decision making for parents at a stressful time [8] and is suitable for comparisons of routine care. Additional opt-in consent was sought later for trial-related imaging procedures. Trial data were obtained from the National Neonatal Research Database (NNRD) [9], supplemented with prospectively recorded data on daily nutritional intake until discharge.

Randomisation was to one of three groups: i) unfortified MM supplemented with unfortified DM (UHM); ii) fortified MM supplemented with fortified DM (FHM); iii) unfortified MM supplemented with preterm formula (PTF). Allocation was computer generated centrally with randomisation to 1 of the 3 feeding groups in a 1:1:1 ratio with minimisation by infant sex, gestational age (25+0-27+6 weeks and 28+0-31+6 weeks), and small for gestational age status (birthweight z score <1.28 and ≥1.28). Twins were randomised individually. Feeding intervention continued until 35+0 weeks postmenstrual age (PMA), after which they were transitioned to suck feeds by breast or bottle according to maternal choice with continuation of any fortification at the discretion of the attending clinician.

Pasteurised (Holder method) donor milk was supplied by the North-West UK Human Milk Bank analysed for macronutrient content pre-pasteurisation [10]. Preterm formula was Cow and Gate Nutriprem 1 (Nutricia Ltd; 80kcal/100ml energy, 2.6g/100ml protein (Non hydrolysed Whey: Casein ratio: 60:40), 3.9g/100ml fat, 8.4g/100ml carbohydrate, see supplementary table S1). Fortifier (Cow and Gate Nutriprem; 15kcal/100ml energy, 1.1g/100ml protein (Extensively hydrolysed Whey:Casein ratio: 50:50), 0g/100ml fat, 2.7g/100ml carbohydrate, see supplementary table S1), was introduced in the FHM group once a total enteral feed volume of 100ml/kg/d was reached.

Milk volumes were increased to between 180-200ml/kg/d for UHM and FHM, and up to 165ml/kg/d for PTF, as per recommendation, modified as necessary in accordance to infant tolerance, and parenteral nutrition weaned in accordance with standard guidelines. Safety criteria defined by slow growth were based on the UK Neonatal and Infant Close Monitoring growth chart 2009 [11, 12]: if after two weeks of reaching a milk volume of 120ml/kg/d, the infant showed a 3 marked centile downward crossing (equating to approximately a 1.4-2.0 z-score change from birthweight) fortification or formula was commenced. All infants were included in the intention-to-treat analysis unless withdrawn.

### Study outcomes

Feasibility outcomes included opt-out, and opt-in consent rates, triggering of safety criteria as described above, adherence to feed intervention, estimation of trial DM requirement, and completeness and accuracy of data retrieval from the NNRD. A conservative 50% consent rate of parents approached was deemed acceptable for both stages. Non-adherence to feed intervention was expected in only a handful of cases. Accuracy and completeness of NNRD baseline clinical characteristic data, and clinical morbidity data (see Table 4) including NEC were assessed by comparison with prospectively collected clinical data from individual patient case notes.

For anthropometry and body composition data, primary outcome was total body adipose tissue (TAT) volume at term (37-42 weeks PMA) as estimated by whole body MRI. Secondary outcomes were i) adipose tissue (AT) depot volumes ii) non-adipose tissue mass (NATM) iii) anthropometry (change in weight, length, and head circumference z score from birth to scan visits), at term and term plus 6 weeks, and iv) TAT at term plus 6 weeks. Post-hoc comparisons between randomised groups of in-hospital change in anthropometry z scores, and macronutrient intake using reference values for MM [13] were also undertaken to aid interpretation of the primary and secondary outcomes.

We scanned infants in accordance with our well-established protocol [14]. TAT volume was quantified as the sum of six discrete depots (Supplementary Fig. 1) as previously described [15]. Image analysis (Slice-OMatic V.4.2; Tomovision) was undertaken independently by Vardis Group, London, UK, blinded to participant identity and feeding group allocation. NATM was calculated as (body weight (g) − (TAT volume (cm3) × 0.9) assuming a value for the density of adipose tissue of 900 g/L. Since there are no reference data informing the impact of preterm diet on adiposity, we used our group’s database of preterm at term body composition scans at the time, which did not have detailed information on diet, and based the sample size on our estimate that 22 infants in each group would provide 80% power (2-sided; 5% significance) to detect a 240cm^3^ (1.0 standard deviation) difference in TAT volume.

### Statistical analysis

All analyses were performed with SPSS version 27. Statistical significance was defined as P<0.05. Differences between unadjusted randomised groups were assessed using ANOVA with examination of the three pairwise differences. To allow for meaningful comparisons of relative adiposity, we adjusted for body size, using ponderal index at term and body mass index (BMI) at term plus 6 weeks [16], PMA at scan, and sex [17]. We used multiple linear regression for adjusted comparisons. We plotted histograms for each outcome, and checked standardised residuals for normality, with natural log transformation of the dependent variable for non-normality. We report the adjusted mean difference with 95% confidence intervals. All analyses were intention to treat. Sensitivity analyses were undertaken for all outcomes for only those that completed their allocated feed intervention, and after excluding outcome outliers (> mean ±3SD).

## Results

We randomised 103 infants, 35 to UHM, 34 to FHM and 34 to PTF groups (Fig. 1). Baseline characteristics were comparable between groups (Table 1).

**Figure 1.**
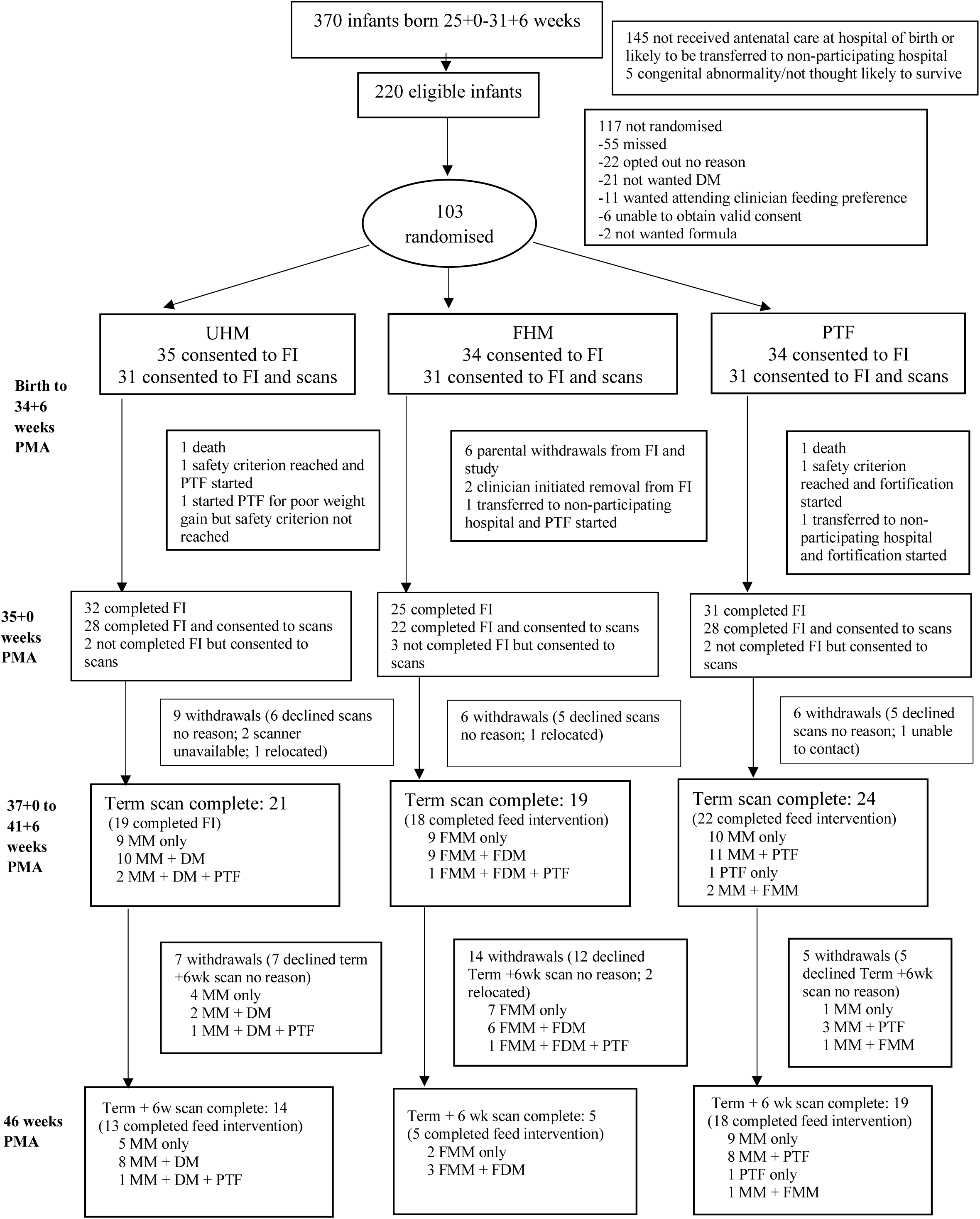
Consort diagram for study infants FI (Feed Intervention Birth to 34+6 weeks PMA)

**Table 1.** Baseline characteristics by randomised feed group.

| Characteristic |  | UHM<br>(n=35) | FHM<br>(n=34) | PTF<br>(n=34) |
| --- | --- | --- | --- | --- |
| Sex, number (%) | Boys | 20 (57) | 21 (62) | 19 (56) |
|  | Girls | 15 (43) | 13 (38) | 15 (44) |
| Gestational age (weeks) at birth, mean (sd) |  | 29.4 (2.2) | 29.6 (2.0) | 29.7 (1.7) |
| Birthweight (g), mean (sd) |  | 1257 (323) | 1312 (333) | 1326 (334) |
| Birthweight z score, mean (sd) |  | -0.42 (0.94) | -0.40 (0.78) | -0.36 (0.88) |
| Birth length (cm), mean (sd) |  | 37.5 (2.9) | 38.2 (3.3) | 38.6 (2.9) |
| Birth length z score, mean (sd) |  | -0.67 (1.15) | -0.60 (1.05) | -0.37 (1.12) |
| Birth OFC (cm), mean (sd) |  | 26.7 (2.8) | 27.2 (2.1) | 27.5 (2.1) |
| Birth OFC z score, mean (sd) |  | -0.77 (1.40) | -0.40 (0.78) | -0.33 (1.03) |
| Multiple birth status, number (%) | Singleton | 17 (49) | 21 (62) | 21 (62) |
|  | Multiple | 18 (51) | 13 (38) | 13 (38) |
| Small for gestational age (Birthweight z score < 1.28), number (%) |  | 6 (17) | 4 (12) | 4 (12) |
| Received antenatal steroids, number (%) |  | 31 (89) | 31 (91) | 33 (97) |
| Apgar score at 5 min, median (IQR) |  | 8.0 (3.0) | 9.0 (1.0) | 9.0 (2.0) |
| Maternal age (years), mean (sd) |  | 33.3 (6.0) | 32.1 (5.6) | 33.4 (6.0) |
| Maternal parity, number (%) | 1 | 28 (80) | 24 (71) | 23 (68) |
|  | >1 | 7 (20) | 10 (29) | 11 (32) |
| Maternal ethnicity, number (%) | White | 25 (71) | 23 (68) | 21 (62) |
|  | Mixed | 3 (9) | 2 (6) | 2 (6) |
|  | Asian | 1 (3) | 1 (3) | 7 (20) |
|  | Black | 5 (14) | 7 (20) | 3 (9) |
|  | Other | 1 (3) | 1 (3) | 1 (3) |

Of 165 parents approached, 103 (62%) chose not to opt-out; 93 out of 165 (56%) consented to both feeding intervention and MRI scans. Reasons for parental opt-out are provided in Figure 1. There were no instances in which the attending clinician felt randomisation was inadvisable. Fifteen infants did not complete feed intervention: there were 6 (6%), parental withdrawals from the study during feed intervention, all from the FHM arm (5 exclusively MM fed; 3 were twins); 5 were removed from feed intervention by the clinician (3 for slow weight gain (2 of which met rescue criterion); 1 with a late diagnosis of duodenal atresia and 1 with feed intolerance); 2 died; and 2 were transferred to a non-participating hospital.

Two infants met the rescue criterion for poor weight gain. One, from the UHM group, received exclusive DM, with a weight z score change from birth to 31+1 weeks of −2.41. The second, from the PTF group received exclusive MM, and had a weight z score change of −1.49 at 32+5 weeks. A third infant (UHM group; received 20% MM, 80% DM) was switched to formula at 34+4 weeks though weight z score change (−1.02) did not reach the pre-specified threshold.

Feed intervention total enteral intake (Supplementary Table S3) and exclusive MM intake (UHM 50%; FHM 44%; PTF 39%) were comparable between groups. The mean volume of DM required to 35+0 weeks PMA for each infant randomised to UHM or FHM groups was 1.3L. There were no significant differences in the volumes of UHM, FHM, and preterm formula from 35 weeks PMA and discharge between the 3 groups. (Supplementary Table S3).

From the UHM, FHM, and PTF groups 21, 19, and 24 infants completed imaging at term, and 14, 5, and 19 respectively, at term plus 6 weeks. There was no significant difference between groups in the unadjusted primary outcome, TAT volume at term (mean (sd): UHM: 0.870L (0.35L); FHM: 0.889L (0.31L); PTF: 0.809L (0.25L), p=0.66), nor in the adjusted TAT (Table 2). We found no significant between group differences for any other body composition measure nor anthropometric measure at term (Table 2 and Fig. 2) or term plus six weeks (Supplementary Tables S2 and Fig. 2).

**Table 2.** Term body composition and anthropometry outcomes by randomised feed group, data are mean (sd) for unadjusted comparisons and mean difference (95%CI) for adjusted comparisons.

| Unadjusted comparisons | Feed group |  |  |  |
| --- | --- | --- | --- | --- |
| Outcome | UHM n*=21 | FHM n*=19 | PTF n*=24 | p# value |
| Δ Wt z score birth to 35 w | -1.20 (0.57) n=34 | -0.79 (0.46) n=28 | -0.79 (0.63) n=33 | <0.01<sup>a</sup> |
| Δ Length z score birth to 35 w | -1.43 (0.73) n=34 | -1.12 (0.68) n=27 | -1.22 (0.70) n=32 | 0.35 |
| Δ OFC z score birth to 35 w | -0.18 (1.02) n=34 | -0.19 (0.76) n=28 | -0.21 (0.98) n=33 | 0.99 |
| PMA (weeks) at Term MRI scan | 40.9 (1.9) | 40.2 (2.1) | 40.9 (1.8) | 0.36 |
| Term MRI weight (g) | 3295 (798) | 3185 (678) | 3153 (656) | 0.79 |
| Term MRI length (cm) | 50.0 (3.8) | 49.5 (3.0) | 49.6 (3.5) | 0.90 |
| Term MRI OFC (cm) | 35.9 (2.4) | 35.0 (1.9) | 35.4 (2.1) | 0.46 |
| Δ Wt z score birth to Term | -0.51 (0.85) | -0.40 (0.75) | -0.75 (0.97) | 0.41 |
| Δ OFC z score birth Term | 1.02 (1.46) n=20 | 0.69 (1.09) | 0.41 (1.15) n=23 | 0.28 |
| Δ length z score birth to Term | -0.49 (1.24) | 0.07 (1.42) | -0.79 (1.18) n=23 | 0.09 |
| TAT at Term (L) | 0.870 (0.35) | 0.889 (0.31) | 0.809 (0.25) | 0.66 |
| Non-ATM at Term (g) | 2511.8 (502.8) | 2385.7 (437.8) | 2424.7 (462.6) | 0.68 |
| %ATM at Term | 23.0 (4.7) | 24.6 (4.5) | 22.8 (3.4) | 0.32 |
| IAAT (L) at Term | 0.043 (0.02) | 0.044 (0.02) | 0.040 (0.01) | 0.67 |
| INAAT (L) at Term | 0.071 (0.03) | 0.072 (0.03) | 0.071 (0.02) | 0.97 |
| DSCAAT (L) at Term | 0.017 (0.01) | 0.017 (0.01) | 0.015 (0.01) | 0.75 |
| DSNAAT (L) at Term | 0.017 (0.01) | 0.017 (0.01) | 0.016 (0.01) | 0.56 |
| SSCAAT (L) at Term | 0.141 (0.07) | 0.137 (0.06) | 0.125 (0.05) | 0.61 |
| SSCNAAT (L) at Term | 0.579 (0.23) | 0.601 (0.21) | 0.543 (0.17) | 0.63 |
| Adjusted Term body composition regression models | Feed group comparison |  |  |  |
| Outcome | PTF (n=24) versus UHM (n=21) (Ref) | FHM (n=19) versus UHM (Ref) | PTF versus FHM (Ref) |  |
| TAT (L)<sup>1</sup> | -0.064 (-0.235 to 0.108), p=0.46 | -0.012 (-0.193 to 0.179), p=0.90 | -0.052 (-0.228 to 0.124), p=0.56 |  |
| TAT (L)<sup>2</sup> | -0.063 (-0.199 to 0.074), p=0.36 | 0.056 (-0.090 to 0.202), p=0.45 | -0.119 (-0.260 to 0.023), p=0.10 |  |
| Non-ATM (g)<sup>1</sup> | -94.0 (-360.4 to 172.5), p=0.48 | -173.8 (-455.9 to 108.3), p=0.22 | 79.9 (-193.1 to 352.8), p=0.56 |  |
| Non-ATM (g)<sup>2</sup> | -89.7 (-282.7 to 103.3), p=0.36 | -65.0 (-271.8 to 141.8), p=0.53 | -24.7 (-225.0 to 175.6), p=0.81 |  |
| % ATM<sup>3</sup> | -0.2 (-2.5 to 2.1), p=0.87 | 2.2 (-0.3 to 4.7), p=0.08 | -2.2 (-4.8 to 0.3), p=0.09 |  |
| IAAT (% difference)<sup>1</sup> | -2.7 (-22.4 to 21.9), p=0.81 | 5.7 (-16.7 to 34.2), p=0.64 | -8.0 (-27.0 to 16.0), p=0.48 |  |
| IAAT (% difference)<sup>2</sup> | -2.5 (-20.2 to 19.1), p=0.80 | 13.0 (-9.0 to 40.1), p=0.26 | -13.7 (-29.9 to 6.3), p=0.16 |  |
| INAAT (L)<sup>1</sup> | -0.002 (-0.018 to 0.015), p=0.84 | -0.003 (-0.021 to 0.015), p=0.73 | 0.001 (-0.016 to 0.018), p=0.88 |  |
| INAAT (L)<sup>2</sup> | -0.002 (-0.016 to 0.012), p=0.82 | 0.003 (-0.012 to 0.018), p=0.70 | -0.004 (-0.019 to 0.010), p=0.54 |  |
| DSCAAT (% difference)<sup>1</sup> | -0.9 (-28.2 to 36.8), p=0.96 | 29.7 (-27.0 to 44.3), p=0.88 | -3.3 (-30.5 to 34.5), p=0.84 |  |
| DSCAAT (% difference)<sup>2</sup> | -0.9 (-24.3 to 29.8), p=0.95 | 15.4 (-13.5 to 54.0), p=0.32 | -14.1 (-35.1 to 13.5), p=0.28 |  |
| DSCNAAT (L)<sup>1</sup> | -0.002 (-0.006 to 0.004), p=0.37 | -0.0002 (-0.004 to 0.004), p=0.92 | -0.002 (-0.005 to 0.002), p=0.44 |  |
| DSCNAAT (L)<sup>2</sup> | -0.002 (-0.005 to 0.002), p=0.32 | 0.001 (-0.003 to 0.004), p=0.65 | -0.003 (-0.006 to 0.001), p=0.16 |  |
| SSCAAT (L)<sup>1</sup> | -0.014 (-0.045 to 0.018), p=0.38 | -0.004 (-0.037 to 0.029), p=0.81 | -0.010 (-0.042 to 0.022), p=0.55 |  |
| SSCAAT (L)<sup>2</sup> | -0.014 (-0.040 to 0.013), p=0.31 | -0.007 (-0.022 to 0.036), p=0.64 | -0.021(-0.048 to 0.007, p=0.14 |  |
| SSCNAAT (L)<sup>1</sup> | -0.041 (-0.156 to 0.073), p=0.47 | -0.004 (-0.125 to 0.118), p=0.95 | -0.038 (-0.155 to 0.080), p=0.52 |  |
| SSCNAAT (L)<sup>2</sup> | -0.041 (-0.132 to 0.050), p=0.37 | -0.041 (-0.056 to 0.139), p=0.40 | -0.082 (-0.177 to 0.012), p=0.09 |  |

\*Number of infants unless otherwise stated; <sup>1</sup> Model Adjusted for Ponderal Index at Term scan (Weight (kg)/Length (M)<sup>3</sup>); <sup>2</sup> Model Adjusted as for model 1 plus Post menstrual age (PMA) at scan, and Sex; <sup>3</sup> Model Adjusted for PMA at scan, and Sex; Wt Weight; OFC Occipito-frontal circumference; TAT Total Adipose Tissue; Non-ATM Non adipose tissue mass; IAAT Internal Abdominal Adipose Tissue; INAAT Internal Non-Abdominal Adipose Tissue; DSCAAT Deep Subcutaneous Abdominal Adipose Tissue; DSCNAAT Deep Subcutaneous Non-Abdominal Adipose Tissue; SSCAAT Superficial Subcutaneous Abdominal Adipose Tissue; SSCNAAT Superficial Subcutaneous Non-abdominal Adipose Tissue; #Between group ANOVA; <sup>a</sup>Tukey post hoc test: PTF>UHM p=0.01; FHM>UHM p=0.01

**Fig. 2.**
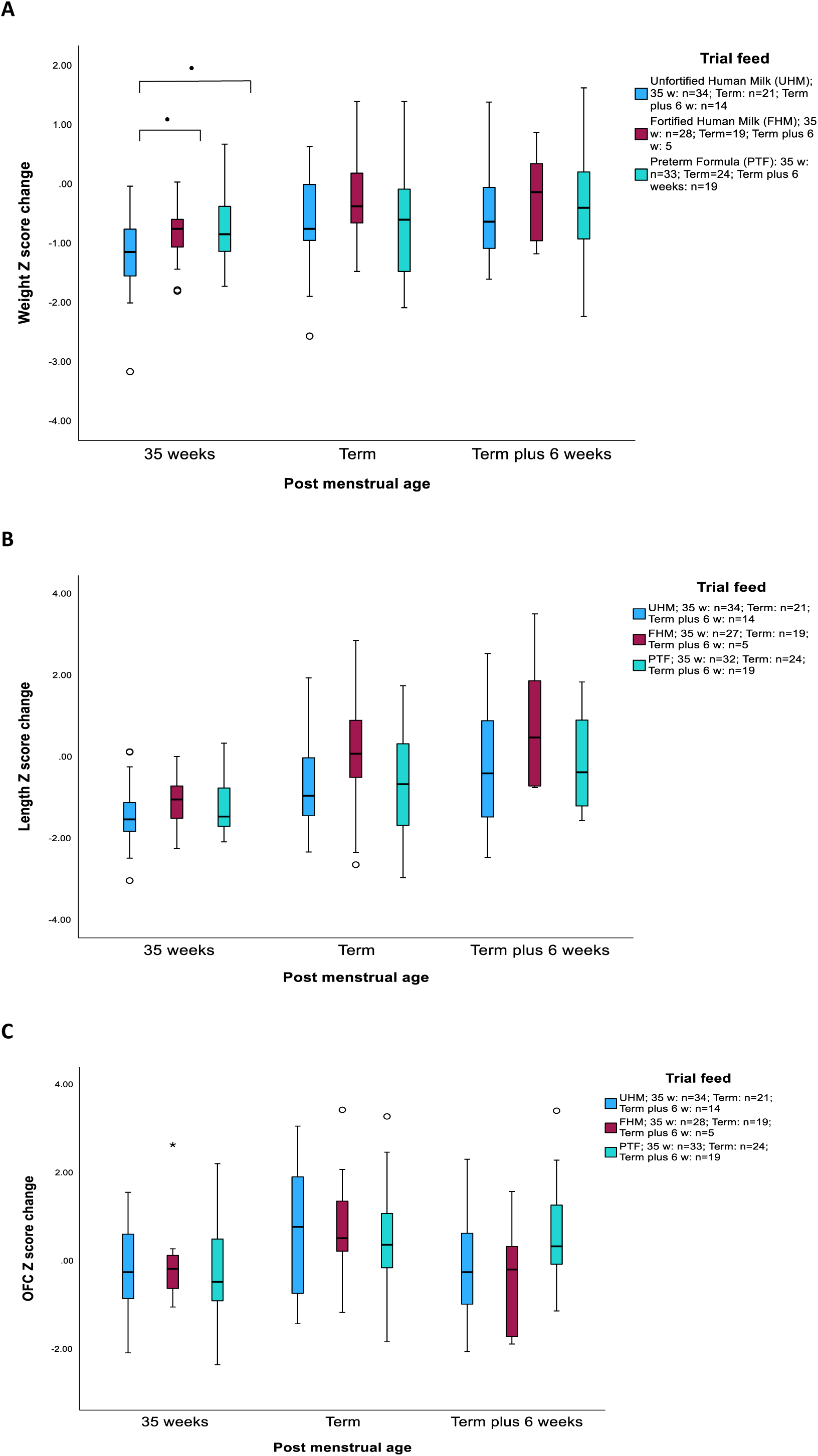
Box plots for (A) weight (B) length, and (C) Occipital Frontal circumference (OFC) Z score change from birth to 35 weeks postmenstrual age, term, and term plus 6 weeks age for randomised feed groups. Solid line is median, box is interquartile range (IQR), whiskers are 1.5 x IQR; ● p <0.05; *Outlier (>1.5 x IQR) ○ Outlier (>3 x IQR).

Post-hoc analyses revealed a fall in weight z score from birth to 35 weeks PMA occurring in all three groups (mean (sd) UHM: −1.20 (0.57); FHM: −0.79 (0.46); PTF −0.79 (0.63)) and was most marked in the UHM group: UHM vs FHM (p=0.01); UHM vs PTF (p=0.01) (Table 2; Fig. 2). Total (parenteral and enteral) mean protein, and protein:energy ratio from birth to 35 weeks PMA, but not energy or fat, were significantly higher in the FHM compared to either UHM and PTF groups (Table 3 and Supplementary Table S3). Sensitivity analyses did not change the body composition, anthropometry, or nutritional intake findings.

**Table 3.** Average daily nutritional intake (parenteral and enteral) for randomised feed groups to 35+0 weeks PMA; data are mean (sd)

| **Outcome** | **UHM<sup>a</sup>** | **FHM<sup>b</sup>** | **PTF<sup>c</sup>** | **p\*$<0.05$** |
| --- | --- | --- | --- | --- |
| Day of age PN stopped | 12 (7) n=35 | 12 (8) n=28 | 11 (7) n=34 |  |
| % MM of total enteral intake | 75.5 (32.1) n=34 | 82.1 (27.5) n=27 | 75.8 (31.3) n=31 |  |
| UMM mLs/kg/d | 90 (47) n=34 | 31 (40) n=27 | 94 (49) n=27 | a>b;c>b |
| UDM mLs/kg/d | 29 (40) n=34 | 4 (8) n=27 | 0 (2) n=28 | a>b;a>c |
| FMM mLs/kg/d | 8 (23) n=34 | 73 (50) n=27 | 3 (10) n=27 | b>a;b>c |
| FDM mLs/kg/d | 0 (0) n=34 | 13 (25) n=27 | 0 (0) n=28 | b>a;b>c |
| PTF mLs/kg/d | 4 (12) n=34 | 3 (9) n=27 | 31 (n=27) | c>a;c>b |
| Total milk mLs/kg/d | 130 (30) n=34 | 124 (34) n=27 | 128 (23) n=27 |  |
| Total milk mLs/kg/d from day reached full enteral feeds<sup>#</sup> | 175 (35) n=34 | 182 (18) n=27 | 170 (23) n=27 |  |
| Protein g/kg/d | 2.8 (0.6) n=34 | 3.6 (0.6) n=27 | 2.9 (0.4) n=29 | b>a;b>c |
| Energy kcal/kg/d | 110 (11) n=34 | 121 (11) n=27 | 109 (9) n=29 | b>a;b>c |
| Pro:En g/100 kcal | 2.5 (0.4) n=34 | 3.0 (0.2) n=27 | 2.6 (0.2) n=29 | b>a;b>c** |
| Fat g/kg/d | 5.0 (0.9) n=34 | 5.1 (0.6) n=27 | 5.1 (0.6) n=29 |  |
| CHO g/kg/d | 13.2 (1.2) n=34 | 15.1 (1.4) n=27 | 12.8 (0.9) n=29 | b>a;b>c |

\*Between group ANOVA post hoc Tukey test, all p-values $<0.001$ except \*\* p $<0.01$ ; <sup>#</sup>Defined as day PN stopped; PN Parenteral Nutrition; DM Pasteurised donor milk (unfortified or fortified); MM Mother's own milk (unfortified or fortified); UMM Unfortified mother's own milk; FMM Fortified mother's own milk; UDM Unfortified pasteurised donor milk; FDM Fortified pasteurised donor milk; PTF Preterm formula; Pro:En Protein:Energy ratio; CHO Carbohydrate

Morbidity and mortality data by feed group are provided in Table 4. There were two deaths (Enterobacter sepsis and suspected sepsis with renal failure, from the UHM and PTF groups respectively), both exclusively MM fed. Two infants, both in the UHM group, developed necrotising enterocolitis (NEC) ≥ Stage II (Bell’s modified criteria). One had surgical NEC and had received MM and DM; the other had NEC stage 2B and received only MM. Baseline characteristics and clinical morbidity data were recorded accurately in the NNRD with 100% agreement between NNRD and prospectively collected data from patient notes. Completeness of data recorded in the NNRD was 100% for clinical morbidity data and the majority of baseline characteristics with only a couple of domains scoring 93% and 94%.

**Table 4.**
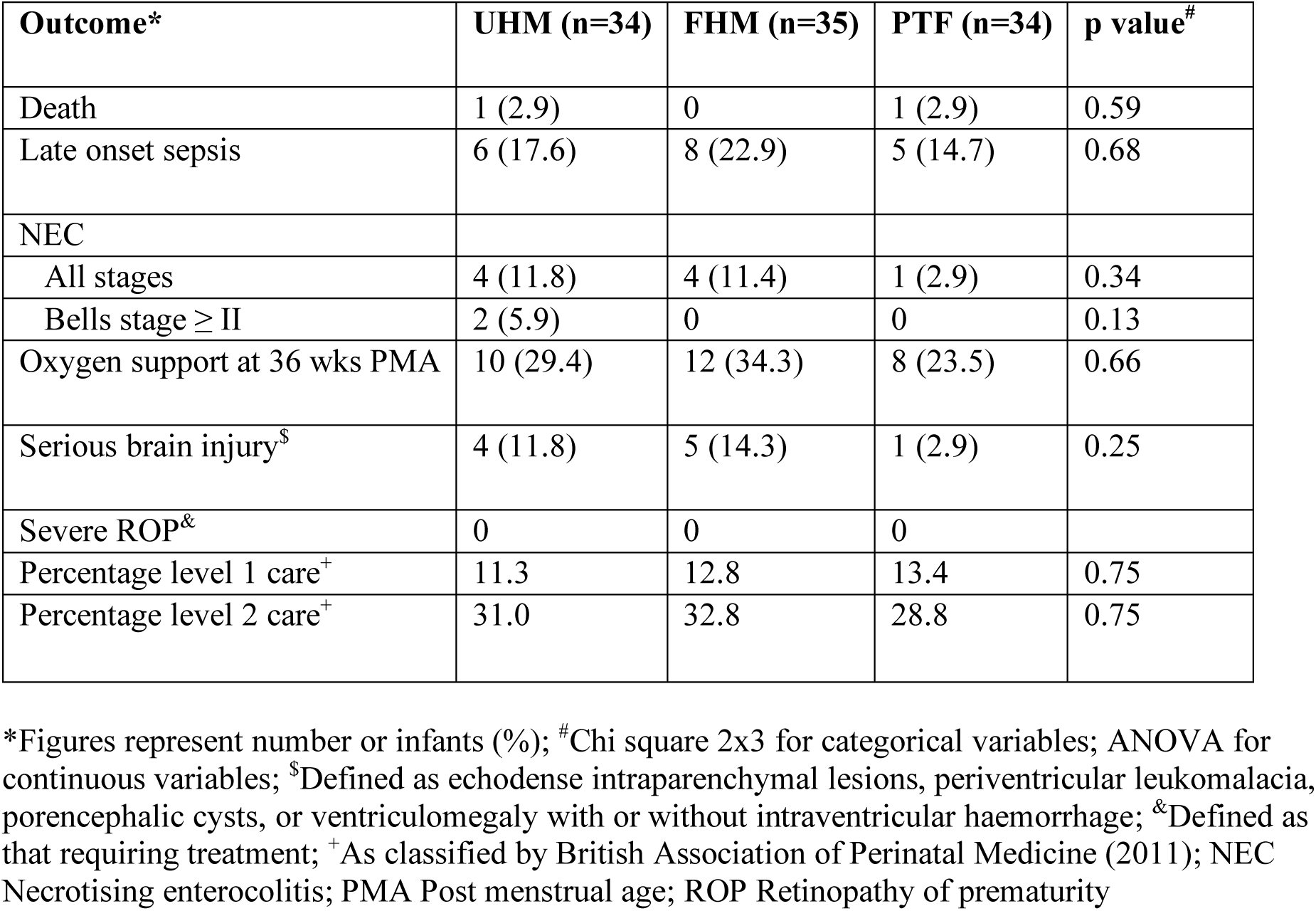
Clinical course by randomised feed groups.

## Discussion

In this very preterm feeding feasibility trial, opt-out consent and randomisation to UHM, FHM, and PTF feeding interventions were acceptable to clinicians and parents, associated with safe growth profiles, with no statistically significant differences in anthropometry or body composition at term or term plus 6 weeks.

As far as we are aware, this is the first RCT to compare these feeding regimens which are in wide use globally. The strengths of the study lie in body composition evaluated using a direct gold-standard approach [18], analyses of imaging data blind to feeding mode, and detailed data on macronutrient intake. Though separating an initial opt-out process from a subsequent opt-in process for non-clinical trial procedures maximises recruitment into all elements of the study, several parents declined the opt-in imaging, resulting in reduction in power for the body composition outcome. Inclusion criteria aimed to minimise loss of study infants from transfer to a non-participating hospital. A major barrier to recruitment was the high number of inborn eligible gestation infants which were not local to the hospital of birth (39%), which may have implications for generalisability.

Feeding intervention consent by opt-out method was 62%, which is higher than recruitment rates for other very preterm nutritional intervention studies using a single opt-in consent process [19, 20].

Using opt out consent increases recruitment rate and generalisability of trial results [21]. There were no instances in which a clinician refused feeding intervention randomisation, and only one in which it was subsequently stopped (fortifier) due to tolerance concerns. We did not anticipate the 6% parental withdrawal rate from the FHM arm. We commenced fortification at 100mls/kg/d, which is routine practice in some countries. However, in the UK, if used, fortification is typically started at a volume of 150ml/kg/d [22]. Early use of fortifier may have introduced bias possibly influencing parent perspectives. Cluster randomisation would be one approach to reducing these biases and has been well received in our parent focus group discussions [23].

All but two of the infants had weight z score changes of no more than −2.0. One of these two infants had surgical NEC, and these data are in keeping with our large UK population cohort study showing weight change at term ranging from −1.3 to −1.8 z-score, and those infants with mortality or serious morbidity having weight z scores up to 1.0 less than those without [24]. There were no statistically significant differences in severe clinical outcomes though the trial was not powered to detect clinically important differences. Data on birth characteristics and clinical course were accurately captured in the NNRD, thus demonstrating the potential for minimising the burden of data capture using linked electronic health care records to achieve long-term follow-up at low cost.

Our finding of no difference in total adiposity is consistent with three previous preterm feeding RCT’s comparing diets of unfortified and fortified MM, fortified DM and PTF as supplements to fortified MM, and unfortified DM and PTF supplements to unfortified MM, finding no difference of fat or fat free mass using different measurement techniques at term [25], 5.5 years [26], and adolescence [27] respectively. We also found no difference in regional adiposity between groups, and taken together, these findings suggest that term age [6] and adult adipose phenotype [5] in individuals born preterm is not mediated by these early diets. Nevertheless, a longer-term programming effect cannot be excluded, and body composition follow up of this cohort is currently underway.

Consistent with significant differences in feeding intervention macronutrient intake, post-hoc analyses revealed that weight gain at the end of the feeding intervention was lower in the UHM than either the PTF or FHM groups, with no significant differences in anthropometry subsequently. This suggests that infants may have self-regulated intake once transitioned to suck feeds, returning to a pre-destined growth trajectory.

Despite higher protein intake in the FHM group compared to UHM or PTF groups during feed intervention, there were no significant differences in NATM at term scan. Observational studies also using reference values for macronutrient content of MM, have associated higher in-hospital protein intakes in very preterm infants, with increased fat free mass at term or near term [28,29], yet with no significant differences between those infants receiving unfortified versus fortified human milk [29]. It is possible that variable content in MM could explain the lack of difference seen in body composition between fortified and unfortified groups. In support of this, we have previously shown in a secondary analysis of very preterm infants randomised to higher and lower protein content in parenteral nutrition [30], that infants predominantly preterm formula fed had higher NATM compared to those exclusively human milk fed. In this study, there was low intake of preterm formula, and a high percent of MM intake across all groups at 75-82%.

In the context of a larger RCT addressing the uncertainties of UHM, FHM, and PTF feeding reliably, powering for NEC as the primary outcome would establish cohorts large enough for long-term follow up health outcomes. We estimate around 14000 infants would be required to identify a 25% relative risk reduction in surgical NEC given current UK baseline rates. An acceptance rate of around 60% as demonstrated in this feasibility study, indicates that the UK, with around 6000 very preterm births each year, could achieve recruitment within a practicable timeframe. From our data we estimate an annual DM requirement for a trial of around 1000L per thousand participants which is readily achievable given total UK human milk bank provision of at least 3500L per annum [31].

In summary, our feasibility data provide justification to proceed to a larger RCT powered to detect differences in important health outcomes.

## Statement of Ethics

*This study protocol was reviewed and approved by the UK National Research Ethics Service, approval number [12/LO/1391]. As detailed in the main body of the text, opt-out and opt-in informed consent protocols were used for use of participant data for research purposes. These consent procedures were reviewed and approved by UK National Research Ethics Service, London – Fulham, approval number [12/LO/1391], 10/12/2012*.

## Author Contributions

NM conceptualized and designed the study, funded the study through unrestricted funds, coordinated and supervised data collection and analyses, and contributed to writing the manuscript, and all revisions. LM drafted and revised study protocol, recruited patients, collected data, carried out analyses, drafted the initial manuscript and reviewed and revised the manuscript. KEC, RE, and AA collected data, carried out the initial analyses and reviewed the manuscript. IA helped in recruitment of patients, collected data, and reviewed the manuscript. RN and JC facilitated recruitment of patients, coordinated data collection, and reviewed the manuscript. SU coordinated and supervised data collection and initial analyses, critically reviewed the manuscript for important intellectual content, reviewed and revised the manuscript. All authors approved the final manuscript as submitted and agree to be accountable for all aspects of the work.

## Data Availability Statement

The data that support the findings of this study are available from the corresponding author, LM, upon reasonable request.

