## Supplemental Tables and Figures for "Preterm formula, fortified or unfortified human milk for very preterm infants, the PREMFOOD study, a parallel randomised feasibility trial"

**Supplementary Tables and Figures**

**Table S1. Macronutrient and mineral composition of Preterm formula (Cow and Gate Nutriprem 1), and Breast milk fortifier (Cow and Gate Nutriprem Breast Milk Fortifier)**

| **Macronutrient and Mineral** | **Cow and Gate Nutriprem 1 per 100ml** | **Cow and Gate Nutriprem Breast Milk Fortifier per 100ml** |
| --- | --- | --- |
| Energy kcal | 80 | 15 |
| Protein g | 2.6 | 1.1 |
| Whey:Casein ratio | 60:40 | 50:50 |
| Carbohydrate g | 8.4 | 2.7 |
| Fat g | 3.9 | 0 |
| Sodium mg | 70 | 35 |
| Potassium mg | 82 | 23 |
| Calcium mg | 101 | 66 |
| Phosphorous mg | 63 | 38 |

**Table S2. Term plus 6 weeks body composition and anthropometry outcomes by randomised feed group, data are mean (sd) for unadjusted comparisons and mean difference (95%CI) for adjusted comparisons**

| **Unadjusted comparisons** | **Feed group** | | | |
| --- | --- | --- | --- | --- |
| **Outcome** | **UHM n=14** | **FHM n=5** | **PTF n=19** | **p^#^ value** |
| PMA (weeks) at T+6w MRI scan | 47.3 (1.2) | 46.7 (1.6) | 47.3 (2.2) | 0.76 |
| T+6w MRI weight (g) | 4564 (982) | 4594 (802) | 4590 (820) | 1.00 |
| T+6w MRI length (cm) | 55.7 (3.5) | 55.9 (2.9) | 56.0 (3.7) | 0.98 |
| T+6w OFC (cm) | 38.8 (2.1) | 37.9 (2.0) | 38.9 (2.0) | 0.49 |
| ∆ Wt z score birth to T+6w | -0.47 (0.70) | -0.18 (1.05) | -0.41 (0.94) | 0.76 |
| ∆ OFC z score birth to T+6w | 0.32 (1.81) | -0.22 (-1.42 to 0.98) | 0.51 (1.25 to 0.29) | 0.53 |
| ∆ length z score birth to T+6w | -0.07 (1.33) | 0.52 (1.87) | -0.17 (1.20) | 0.51 |
| TAT at T+6w (L) | 1.355 (0.486) | 1.474 (0.463) | 1.353 (0.284) | 0.76 |
| Non-ATM at T+6w (g) | 3264.0 (609.9) | 3208.1 (526.2) | 3364.4 (634.1) | 0.82 |
| %ATM at T+6w | 26.6 (5.5) | 28.9 (5.0) | 26.5 (3.7) | 0.48 |
| IAAT (L) at T+6w | 0.064 (0.023) | 0.061 (0.025) | 0.062 (0.018) | 0.97 |
| INAAT (L) at T+6w | 0.109 (0.039) | 0.110 (0.036) | 0.114 (0.033) | 0.91 |
| DSCAAT (L) at T+6w | 0.034 (0.023) | 0.031 (0.028) | 0.031 (0.011) | 0.91 |
| DSNAAT (L) at T+6w | 0.019 (0.009) | 0.024 (0.009) | 0.025 (0.008) | 0.25 |
| SSCAAT (L) at T+6w | 0.223 (0.103) | 0.253 (0.095) | 0.224 (0.053) | 0.67 |
| SSCNAAT (L) at T+6w | 0.907 (0.306) | 0.994 (0.299) | 0.898 (0.193) | 0.68 |
| **Adjusted Term plus 6w body composition regression models** | **Feed group comparison** | | | |
| **Outcome** | **PTF (n=19) versus UHM (n=14) (Ref)** | **FHM (n=5) versus UHM (Ref)** | **PTF versus FHM (Ref)** | |
| TAT (L)^1^ | -0.029 (-0.245 to 0.188), p=0.79 | 0.108 (-0.171 to 0.387), p=0.44 | -0.136 (-0.396 to 0.123), p=0.29 | |
| TAT (L)^2^ | -0.052 (-0.246 to 0.143), p=0.59 | 0.141 (-0.111 to 0.393), p=0.26 | -0.193 (-0.428 to 0.043), p=0.11 | |
| Non-ATM (g)^1^ | 56.1 (-258.9 to 371.1), p=0.72 | -74.8 (-480.7 to 331.2), p=0.71 | 130.9 (-246.6 to 508.3), p=0.49 | |
| Non-ATM (g)^2^ | 6.0 (-267.6.0 to 279.5), p=0.97 | -4.9 (-359.5.0 to 349.8), p=0.98 | 10.8 (-320.5 to 342.1), p=0.95 | |
| % ATM^3^ | -0.04 (-3.4 to 3.4), p=0.98 | 2.6 (-1.8 to 7.0), p=0.23 | -2.7 (-6.8 to 1.5), p=0.20 | |
| IAAT (L)^1^ | -0.003 (-0.019 to 0.014), p=0.75 | -0.002 (-0.023 to 0.019), p=0.82 | 0.0001 (-0.020 to 0.019), p=0.99 | |
| IAAT (L)^2^ | -0.004 (-0.020 to 0.012), p=0.65 | -0.0004 (-0.021 to 0.020), p=0.97 | -0.003 (-0.023 to 0.016), p=0.74 | |
| INAAT (L)^1^ | 0.003 (-0.018 to 0.025), p=0.76 | 0.0002 (-0.027 to 0.028), p=0.99 | 0.003 (-0.022 to 0.029), p=0.81 | |
| INAAT (L) ^2^ | 0.001 (-0.020 to 0.021), p=0.94 | 0.003 (-0.023 to 0.030), p=0.81 | -0.002 (-0.027 to 0.022), p=0.85 | |
| DSCAAT (% difference)^1^ | 4.4 (-33.9 to 64.5), p=0.85 | -22.5 (-56.9 to 39.5), p=0.38 | 34.5 (-22.1 to 132.3), p=0.28 | |
| DSCAAT (% difference)^2^ | 2.7 (-33.3 to 58.2), p=0.90 | -19.8 (-54.3 to 40.4), p=0.43 | 28.1 (-24.0 to 116.2), p=0.34 | |
| DSCNAAT (L)^1^ | 0.004 (-0.04 to 0.012), p=0.29 | 0.005 (-0.001 to 0.011), p=0.11 | 0.001 (-0.007 to 0.008), p=0.82 | |
| DSCNAAT (L)^2^ | 0.005 (-0.001 to 0.011), p=0.13 | 0.005 (-0.003 to 0.013), p=0.21 | -0.0003 (-0.008 to 0.007), p=0.93 | |
| SSCAAT (L)^1^ | 0.028 (-0.032 to 0.089), p=0.35 | -0.004 (-0.051 to 0.043), p=0.87 | -0.032 (-0.089 to 0.024), p=0.26 | |
| SSCAAT (L)^2^ | -0.007 (-0.049 to 0.035), p=0.73 | 0.037 (-0.017 to 0.091), p=0.18 | -0.044 (-0.095 to 0.07), p=0.09 | |
| SSCNAAT (L)^1^ | -0.027 (-0.166 to 0.113), p=0.70 | 0.080 (-0.099 to 0.260), p=0.37 | -0.107 (-0.274 to 0.060), p=0.20 | |
| SSCNAAT (L)^2^ | -0.042 (-0.167 to 0.084), p=0.51 | 0.098 (-0.065 to 0.261), p=0.23 | -0.140 (-0.292 to 0.013), p=0.07 | |

^1^ Model Adjusted for Body Mass Index at Term plus 6 weeks scan (Weight (kg)/Length (M)^2^); ^2^ Model Adjusted as for model 1 plus Post menstrual age (PMA) at scan, and Sex; ^3^ Model Adjusted for PMA at scan and Sex;T+6w Term plus 6 weeks; Wt Weight; OFC Occipito-frontal circumference; TAT Total adipose tissue; Non-ATM Non Adipose Tissue Mass; IAAT Internal Abdominal Adipose Tissue; INAAT Internal Non-Abdominal Adipose Tissue; DSCAAT Deep Subcutaneous Abdominal Adipose Tissue; DSCNAAT Deep Subcutaneous Non-Abdominal Adipose Tissue; SSCAAT Superficial Subcutaneous Abdominal Adipose Tissue; SSCNAAT Superficial Subcutaneous Non-abdominal Adipose Tissue; ^#^Between group ANOVA

**Table S3. Average daily milk and total macronutrient (parenteral and enteral^$^) intake, and or anthropometric change between birth and 35+0 weeks PMA (end of feed intervention) (T1), 35 weeks PMA and discharge (T2), birth and discharge (T3), time from regained birthweight and 35 weeks PMA (T4), 35 weeks PMA and term equivalent age (T5), and birth and term equivalent age (T6), by randomised feed group***

| **Outcome** | **UHM^a^** | **FHM^b^** | **PTF^c^** | **p^#^<0.001** | **p^#^<0.01** | **p^#^<0.05** |
| --- | --- | --- | --- | --- | --- | --- |
| UMM T1 mLs/kg/d | 90 (47) n=34 | 31 (40) n=27 | 94 (49) n=27 | a>b;c>b |  |  |
| UMM T2 mLs/kg/d | 85 (69) n=32 | 85 (63) n=26 | 95 (66) n=26 |  |  |  |
| UMM T3 mLs/kg/d | 89 (48) n=34 | 47 (37) n=27 | 97 (47) n=29 | c>b | a>b |  |
| UDM T1 mLs/kg/d | 29 (40) n=34 | 4 (8) n=27 | 0 (2) n=28 | a>b;a>c |  |  |
| UDM T2 mLs/kg/d | 2 (8) n=32 | 0 (0) n=26 | 0 (0) n=26 |  |  |  |
| FMM T1 mLs/kg/d | 8 (23) n=34 | 73 (50) n=27 | 3 (10) n=27 | b>a;b>c |  |  |
| FMM T2 mLs/kg/d | 11 (39) n=32 | 26 (43) n=26 | 5 (24) n=26 |  |  |  |
| FMM T3 mLs/kg/d | 9 (22) n=34 | 63 (41) n=27 | 3 (10) n=29 | b>a;b>c |  |  |
| FDM T1 mLs/kg/d | 0 (0) n=34 | 13 (25) n=27 | 0 (0) n=28 | b>a;b>c |  |  |
| FDM T2 mLs/kg/d | 0 (0) | 2 (6) n=26 | 0 (0) n=26 |  |  |  |
| Form T1 mLs/kg/d | 4 (12) n=34 | 3 (9) n=27 | 31 (n=27) | c>a;c>b |  |  |
| Form T2 mLs/kg/d | 52 (64) n=32 | 36 (60) n=26 | 44 (59) n=26 |  |  |  |
| Form T3 mLs/kg/d | 16 (24) n=34 | 14 (25) n=27 | 35 (44) n=29 |  |  | c>a;c>b |
| Total milk T1 mLs/kg/d | 130 (30) n=34 | 124 (34) n=27 | 128 (23) n=27 |  |  |  |
| Total milk T2 mLs/kg/d | 150 (48) n=32 | 149 (35) n=26 | 144 (47) n=26 |  |  |  |
| Protein T1 g/kg/d | 2.8 (0.6) n=34 | 3.6 (0.6) n=27 | 2.9 (0.4) n=29 | b>a;b>c |  |  |
| Protein T2 g/kg/d | 2.7 (1.2) n=33 | 2.8 (0.9) n=27 | 2.5 (1.0) n=28 |  |  |  |
| Protein T3 g/kg/d | 2.8 (0.6) n=34 | 3.4 (0.5) n=27 | 2.8 (0.4) n=29 | b>a |  | b>c |
| Energy T1 kcal/kg/d | 110 (11) n=34 | 121 (11) n=27 | 109 (9) n=29 | b>a;b>c |  |  |
| Energy T2 kcal/kg/d | 102 (39) n=33 | 106 (27) n=27 | 99 (37) n=28 |  |  |  |
| Energy T3 kcal/kg/d | 110 (12) n=34 | 116 (12) n=27 | 109 (11) n=29 |  |  | b>c |
| Pro:En T1 g/100 kcal | 2.5 (0.4) n=34 | 3.0 (0.2) n=27 | 2.6 (0.2) n=29 | b>a | b>c |  |
| Pro:En T2 g/100 kcal | 2.6 (0.3) n=33 | 2.6 (0.3) n=27 | 2.5 (0.2) n=28 |  |  |  |
| Pro:En T3 g/100 kcal | 2.5 (0.4) n=34 | 2.9 (0.2) n=27 | 2.6 (0.2) n=29 | b>a | b>c |  |
| Fat T1 g/kg/d | 5.0 (0.9) n=34 | 5.1 (0.6) n=27 | 5.1 (0.6) n=29 |  |  |  |
| Fat T2 g/kg/d | 5.1 (1.9) n=33 | 5.3 (1.3) n=27 | 5.1 (1.9) n=28 |  |  |  |
| Fat T3 g/kg/d | 5.1 (0.8) n=34 | 5.2 (0.6) n=27 | 5.2 (0.6) n=29 |  |  |  |
| CHO T1 g/kg/d | 13.2 (1.2) n=34 | 15.1 (1.4) n=27 | 12.8 (0.9) n=29 | b>a;b>c |  |  |
| CHO T2 g/kg/d | 11.1 (4.3) n=33 | 11.7 (3.0) n=27 | 10.8 (4.00) n=28 |  |  |  |
| CHO T3 g/kg/d | 12.9 (1.28) n=34 | 14.0 (1.6) n=27 | 12.5 (1.2) n=29 | b>c | b>a |  |
| Wt gain T1 g/kg/d^+^ | 9.4 (2.9) n=34 | 11.0 (2.6) n=28 | 11.4(2.8) n=33 |  |  | c>a |
| Wt gain T2 g/kg/d^+^ | 14.7 (6.4) n=32 | 11.6 (4.2) n=26 | 10.8 (3.9) n=30 |  |  | a>c |
| Wt gain T3 g/kg/d^+^ | 10.5 (2.6) n=34 | 11.5 (2.0) n=28 | 11.2 (2.5) n=33 |  |  |  |
| Wt gain T4 g/kg/d^+^ | 12.9 (4.1) n=33 | 14.6 (3.9) n=26 | 14.88 (4.0) n=29 |  |  |  |
| Wt gain T5 g/kg/d^+^ | 12.8 (2.9) n=21 | 12.8 (3.6) n=19 | 10.8 (2.5) n=24 |  |  |  |
| ∆ Wt z score T5 | 0.52 (0.81) n=21 | 0.38 (0.73) n=19 | 0.02 (0.61) n=24 |  |  |  |
| OFC gain T1 cm/wk | 0.7 (0.3) n=34 | 0.7 (0.2) n=28 | 0.7 (0.3) n=33 |  |  |  |
| OFC gain T6 cm/wk | 0.8 (0.2) n=21 | 0.8 (0.1) n=19 | 0.7 (0.1) n=24 |  |  |  |
| Length gain T1 cm/wk | 0.9 (0.2) n=34 | 0.9 (0.3 n=28 | 0.9 (0.3) n=33 |  |  |  |
| Length gain T6 cm/wk | 1.0 (0.2) n=21 | 1.1 (0.3) n=19 | 1.0 (0.2) n=24 |  |  |  |

*Data are presented as mean (sd); ^#^Between group ANOVA post hoc Tukey tests; ^$^Enteral intake defined as all naso-gastric/oro-gastric or bottle milk feeds – ie breastfeeding excluded; ^+^ Defined as ((1000 x ln(Wn/W1))/(Dn-D1) where Wn is weight on n days from Weight 1 (W1) and Dn-D1 is number of days from W1 to Wn.;UMM Unfortified Maternal Milk; FMM Fortified Maternal Milk; UDM Unfortified pasteurised donor milk; FDM Fortified pasteurised donor milk; Form Formula (Preterm or Other); Pro:En Protein:Energy ratio; CHO Carbohydrate; Wt Weight; OFC Occipitofrontal circumference

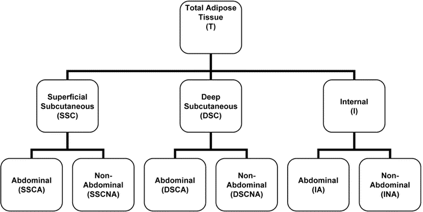

**Figure S1. Classification of adipose tissue depots. Reproduced with permission (ref 16)**
